# Strategies for antigen testing: An alternative approach to widespread PCR testing

**DOI:** 10.1101/2021.04.05.21254024

**Authors:** Sanjay G. Reddy, Saumya Das

## Abstract

Multiple applications of low cost and rapid antigen tests for individuals, in parallel (at the same time) or at times sufficiently close to each other, when appropriately interpreted can considerably increase sensitivity of these tests, improving on their performance greatly. Under reasonable assumptions, this occurs when considering a positive to arise in a composite test if at least one of two underlying repeated tests are positive. Parallel Rapid Testing can potentially provide a form of testing that is accessible (combining wide availability and lack of expense) and quick. Moreover, it can provide a level of sensitivity that is comparable to seemingly more sophisticated but more expensive alternatives. In combination with sequential testing, this strategy offers an alternative method of testing that can be applied immediately and on a widespread basis.

## I. Introduction: The need for accessible, rapid and sensitive tests

Why is testing not more widespread as a strategy for controlling the spread of Covid-19 (by sequestering those who test positive and allowing those who test negative to go about their daily activities)?^2^ Even with the roll-out of vaccination, there is reason to believe that a protracted time interval may be needed to achieve a sufficient level of population immunity to contain the pandemic in many countries, and globally. The emergence of novel strains threatens to extend that timeline further, and therefore the identification and isolation of infected individuals remains imperative. The ‘gold standard’ tests currently used are PCR tests that detect viral RNA in biospecimens collected from nasopharyngeal swabs and have extremely high sensitivity for detecting viral RNA. The best in class of these assays have a limit of detection (LOD) of 100 copies/ml^1^, although there remains significant variation in the performance of the multiple PCR tests that are used under Emergency Use Authorization. The apparently extremely high sensitivity (and hence low rate of false negatives for viral loads above the LOD), is the primary reason these tests have been considered superior. However, there are several drawbacks to PCR tests. These tests are expensive and often only produce results with delays of up to several days (depending on the testing site) that make them all but unusable for the purpose of determining whether tested individuals may be infectious at the time of testing. This limits the ability to control the spread of the disease by restricting the contacts of infected individuals, resulting in massive social and economic costs. Secondly, the extremely high analytic sensitivity of these tests allows for detection of viral loads that may not be associated with infectivity^2^. Mina and colleagues have noted that a significant proportion of PCR positive tests correspond to the long ‘tail’ of viral load in the nasopharynx in the post-infectious patient, and in low prevalence settings, the higher number of ‘false positives’ that may not be clinically relevant^3^ or infective may unnecessarily delay the reopening of institutions such as schools. Alternative test strategies that can identify the infectious or ‘pre-infectious’ patients in a rapid fashion would provide far more use in the general population, where a large majority of patients continue to be asymptomatic even when infectious. If these tests were cheap and quick, society could resume many activities very quickly, even in the absence of mass vaccination. Yet few if any societies have succeeded in launching testing on the scale that many have advocated^4 5^. Insofar as widespread vaccination is only now being undertaken, and very unevenly across countries, complementing vaccination with widespread rapid testing can still aid disease control and accelerate normalization, with attendant social and economic benefits. The strategy we propose here can potentially also be applied to testing strategies for control of other existing and future diseases.

Rapid antigen testing using a variety of platforms was developed to address these needs. However, there has so far not been wide-spread implementation of these tests. The primary reason for the limited adoption of widespread antigen testing lies in the decreased sensitivity of this test when compared to the PCR test. A recent study^6^ evaluating the performance of the Abbott BinaxNOW COVID-19 rapid antigen test with the RT-PCR test to determine the LOD, demonstrated that the threshold of detectability of the Binax-Cov2 test was between 1.6-4.3 × 10^4^ viral copies as opposed to 100 copies for the RT-PCR test. In terms of the more widely used CT (cycle threshold) values, this corresponded to a CT value of 30.3-28.8. In contrast, the RT-PCR test is designated as ‘positive’ if CT < 45 (or 2^15^ fold fewer viral copies). Using a conservative CT value of < 33 to identify ‘true or clinically relevant’ positives and negatives, the sensitivity of the Binax-CoV2 test was 93.8% (CI: 69.8%-99.8%), while the specificity was almost 100% (CI: 99.6-100%). This range of estimates has been replicated in multiple other studies, which have found the sensitivity of antigen tests from several manufacturers to range from 77% to 89% when assessed across the whole range of RT-PCR positive subjects, while the specificity remains close to 100%^7,8^. When restricting to samples obtained from either symptomatic patients or samples with CT < 32, which appear to best correlate with a viral load that may be sufficient to be infective, the sensitivity increases to 95% or higher in some studies, although other studies have suggested a sensitivity > 92.3% only in patients with > 10^6^ viral particles/swab^9^. Nonetheless, the decreased sensitivity of the antigen tests has led to the concern that false negative tests may allow for infective individuals to further expose other individuals thereby leading to propagation of infection. Such concerns have been amplified by several highly publicized ‘super-spreader’ events (such as a White House party to celebrate the nomination of Judge Amy Coney Barrett).

What are the reasons for the lower sensitivity of the rapid antigen test? As noted above, the antigen tests do not apply any amplification step to the input material, and hence have a lower LOD compared to the RT-PCR test. Moreover, most rapid antigen tests are based on lateral flow chromatography and provide binary positive or negative results (with the threshold that separates these results depending on strength and size of the colorimetric band). Thus, the coefficient of variation (CV) of the test can be considerable, especially in samples that are on the lower spectrum of viral load, leading to variability of interpretation of the same test^10^ or of replicates drawn from the same subject^11^. Few studies have rigorously investigated the contribution of variation arising from distinct sources to the overall variability of test results^12^ (such as differences in operator characteristics, in test interpretation, and in technical properties of the test itself).

Secondly, the clinical significance of patients who test negative according to a rapid antigen test but have a positive PCR test (namely patients with CT values in the 28-45 range, or viral loads between 43000 and 100 copies^1^) is important to consider. A retrospective study from hospitalized patients^13^ determined the correlation of RT-PCR CT value on clinical samples with viral infectivity as assessed by infection of cultured VeroE6 cells. In this study, viral infectivity corresponded to a CT value of 18.8 +/- 3.4; samples with a CT value < 23 yielding 91.5% of the isolates; viral infectivity was associated with relatively high viral loads. A second study from symptomatic patients similarly demonstrated a higher correlation between viral infectivity and antigen test positivity than with RT-PCR positivity at prevalently applied CT values^14^. Finally, a recent study recapitulated these findings, demonstrating low probability of infectivity at CT values > 24^15^. These and other studies create a strong case for the hypothesis that viral loads > 10^6^ particles or CT values <24 (as a conservative estimate)^16^,^17^, correlate well with infectivity and are also above the LOD for rapid antigen tests (see **Figure 1:** solid line for clinically relevant positive test, likely corresponding to CT values at or below 24; dashed line depicting LOD for antigen tests and corresponding to CT values of 29-30^18^).

**Figure 1.**
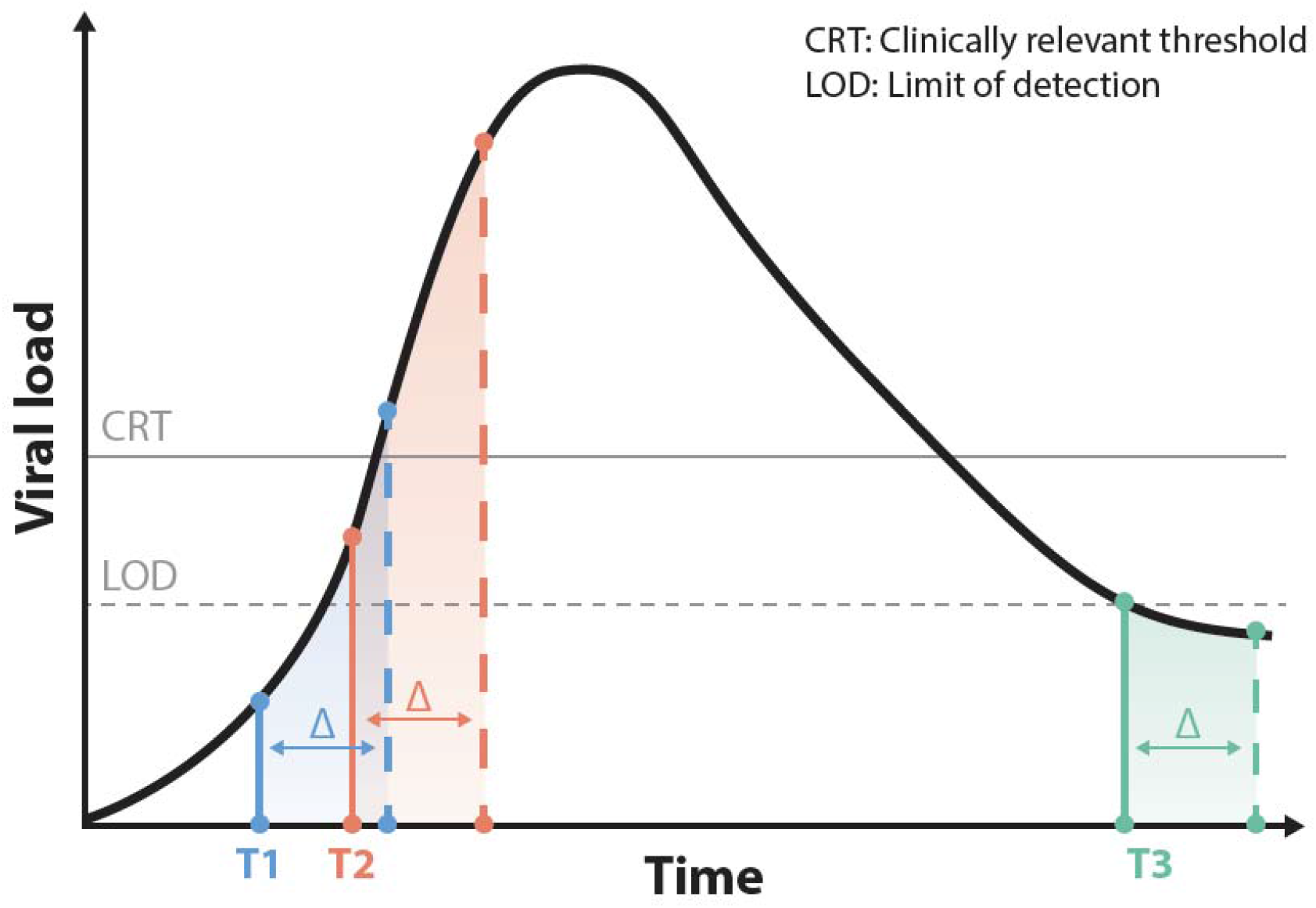
Impact of serial or parallel testing strategy on detection of Sars-Cov2 at different time points along disease trajectory. The graph depicts viral load as a function of time (not to scale). The long ‘tail’ after recovery from clinical infection has been previously well described and therefore only the first part of this tail is depicted. The horizontal solid line depicts a clinically relevant threshold: for instance, a viral load corresponding to this level or higher may correlate with infectivity at that time. This level of viral load will also generate a likely positive test result in a rapid antigen test, as it is above the limit of detection (LOD) depicted in the dashed line. The threshold of clinical relevance is shown as being above the LOD but this is purely illustrative. Because of variability in test performance, technical replicates of samples may lead to different results on repeated testing. For the patient tested at time T1, as the viral load is below the LOD, parallel repeat testing is likely to yield repeated negative results. A patient tested at time T2 may be identified as negative on the basis of a single test, but with parallel testing, replicates are likely to lead to an overall positive test result, hence identifying the patient as a likely infectious patient. For both T1 and T2, serial repeat testing after an interval (Δ) would be still more likely to yield positive tests as the likelihood of a patient testing negative is likely to decrease as the viral load increases. For a patient on the tail end of infection at or beyond Time T3, negative test results are likely to arise for both strategies, but as such a patient is unlikely to be infective, this result is of lesser concern. The figure illustrates the role that both serial and parallel testing can play in clinical management and public health strategies.

Thus, the antigen-test negative, but PCR-test positive patients fall into three categories: i) those patients who have passed their peak of infection and may have detectable viral RNAs in their nasopharynx but rarely have a viral load sufficient to transmit infection (corresponding to time T3 in Figure 1); ii) patients who are ‘pre-symptomatic’ or ‘pre-infectious’ whose viral load at the time of testing is below the LOD for the antigen test, but not the PCR test (time T1, Figure 1), and iii) patients whose viral loads are at least as high as the LOD of the antigen-test, but are interpreted by it as negative due to test variability (time T2, Figure 1). The latter two classes of individuals are of greatest concern, as its members may be infective either at the time of testing or in the following days. There may therefore be particular reason to worry that tests with a higher LOD or lower level of sensitivity than a PCR test may not pick up such individuals in these two groups.

One strategy that has been proposed to address the problem of potential infectiousness of patients with negative antigen-test results is that of serial testing^19^. Paltiel et al, have demonstrated that frequent testing of the population even with a test with sensitivity in the 70% range can control the spread of COVID infection. As discussed in Appendix 1, serial testing is likely to help to identify patients at time points T1 and T2 in Figure 1 -who may either become infective or already be infective – but may not initially generate a positive test. Here we present a complementary strategy to improve the sensitivity of antigen tests and enhance their usefulness in clinical and public health applications, especially relevant for samples that fall in scenario iii) above, who have viral loads above the LOD of an antigen test but are not initially detected. This approach aims to take advantage of the accessibility – widespread availability as well as low cost - and rapid test results of antigen tests but enhance their sensitivity. The method of Parallel Rapid Testing or PRT applies antigen tests multiple times on biological replicate samples and analyzes their results jointly. We shall show that such an approach leads to substantially more accurate results while maintaining reasonable costs. It does so by raising the sensitivity of tests when it most matters, namely because of the possibility that erroneous results (false negatives) can arise for individuals who possess viral loads that have clinical or public health relevance. The benefit of better “disambiguating” false from true negatives will be greatest when sensitivity of an underlying test is low enough to create sizable potential gains from repetition.

Suppose that Test 1 (e.g. a PCR test) has sensitivity of p1, and specificity of q1, and that Test 2 (e.g. an antigen test) has sensitivity of p2 and specificity of q2, with p2< p1 and q2<=q1. Despite the lower sensitivity of Test 2, its use might still be defended on the basis of a tradeoff between cost and speed on the one hand and accuracy on the other. But if there were another test that has the same or higher sensitivity and similar or higher specificity as compared to Test 1 while producing results at greater speed and at lower cost with more clinically meaningful results, there would be no need for a tradeoff at all. In that case, this test - call it Test 3 -would now be a “dominant” test in the sense of being superior according to *all* criteria.

Based on current information about the properties of the available tests, there is indeed such a Test 3, which comes *simply* from repeating test 2 for the same individual – not every few days or every week as has been previously suggested^20^-but immediately. Test 3 involves simultaneously running antigen-tests from biological replicates (separate swab samples) for the same individual and interpreting the resulting *ensemble* of test results appropriately. We call a test formed in this way a “Parallel Rapid Test” (PRT).

How an ensemble of results should be treated will depend on whether it is considered more important to avoid “false positives” or “false negatives”. For instance, if it is deemed most important to avoid false negatives, so as not to provide a license to participate in societal activities to persons who may in fact be infected (and to successfully isolate all of those with the condition) then it might be required that all test results be negative for it to be considered that Test 3 (the repeated test) is “negative”. i.e. a single positive result in the ensemble would be sufficient to lead the composite test based on repetition to be considered “positive”. Such a determination need not be final. Individuals can be isolated and retested the subsequent days with another antigen test and/or monitored for symptoms. Considering a single positive in the ensemble to make for a positive overall in the composite test is only one possible choice. Which approach to take to mixed results depends on the statistical “loss function” of the decision maker -considering the cost of testing and weighing false positives against false negatives -which must be informed by clinical and public health objectives. A decision rule that is more permissive in determining whether mixed test results count as a “positive” will, for a fixed number of tests in an ensemble, necessarily increase sensitivity at the cost of decreasing specificity. Since leading antigen tests appear already to have extremely high specificity, this tradeoff is negligible, however. We show below that, for antigen testing for Covid-19, PRT therefore offers an attractive way to improve greatly overall test performance. By combining the accessibility and rapidity of results provided by antigen tests with the increased sensitivity provided by parallel testing, this approach can be especially useful in identifying infections in the class of patients in group iii) who may conservatively be considered to be potentially infective at the time of testing or shortly thereafter.

## II. A Simple Example of Parallel Rapid Testing

The specificity and sensitivity of a test can only properly be defined on the basis of a specific understanding of what it means to have or not to have the underlying condition being tested (a “true positive”). For instance, the chance that a test incorrectly returns the result that the viral load in an individual is below a specified threshold -when it is in fact above that threshold -determines its sensitivity.

The thresholds of viral load used to define the sensitivity and specificity of a test that is applied for decision-making ought to be influenced by considerations of clinical and public health relevance. In principle, these thresholds may be below or above a test’s LOD. For a binary test, if the clinically relevant threshold for determining the presence of a condition is beneath the test’s LOD, then a viral load above the clinically relevant threshold but below the LOD will be undetectable, and thus limit the test’s sensitivity. If the clinically relevant threshold for determining the presence of a condition is above a binary test’s LOD, then a viral load below the clinically relevant threshold but above the LOD will be detectable, and thus limit the test’s specificity. It is also important to keep in mind that test variability (which as noted above may be affected by multiple factors) can influence whether a sample from a patient tests positive or negative. Especially for viral loads nearer to the LOD, technical replicates, or biological replicates from the same patient may vary in terms of test results. Thus, both the sensitivity and specificity of a test may depend on the level of viral load, because of the role of the LOD in determining whether viral concentrations are detectable at all, and because of the further influence of viral load on the probability of a positive or a negative test. The chance of a ‘false negative’ may decrease (and the sensitivity of a test correspondingly increase) as the viral load in the individual rises, making it easier to identify a clinically relevant viral load, especially once it is above the LOD.

While recognizing these considerations, we focus here – with generality -on a clinical situation in which an individual possesses an unknown viral load, for which a test applied even a single time has associated sensitivity and specificity. We consider the mathematical basis for inference based on parallel testing (immediate or nearly immediate repetition) in such a case. PRT will be especially useful at a time point such as T2 (**Fig. 1**) during the course of an infection, where the viral load is above the LOD for the test, rather than T1, where the viral load is below the LOD for the test, since the likelihood that the patient will return a positive result in any of the tests when the viral load is below the LOD is negligible or zero. The mathematical argument we present applies to all cases, however, since there is a possibility of false positives and of false negatives in any scenario, and that is all that is required to study the inferential use of test repetition.

Consider initially the case in which a single positive result in a repeated test suffices for the composite test to be considered positive. Suppose that Test 2 provides results in a few minutes (as antigen tests can) and costs $c2 per test, as compared with Test 1, which provides results in a number of days but costs $c1 per test. Then Test 3(M) – repeating Test 2 M times -has a total cost of $Mc2. However, the sensitivity of Test 3(M) is p3 (M) = 1-(1-p2)^M^, as long as the probability of a false negative in each test is identical and independent -unaffected by the results of other tests in the ensemble of repeated tests. This is most likely to be the case if distinct biological replicate samples are taken from the individual and if separate tests are applied to each sample. The assumption that the probability of an erroneous result is identical and independent across repeated tests amounts to holding that the *distribution* of test statistics (the likelihood of a correct or erroneous result) does not vary from round to round during immediate repetitions. Note that this distribution may be specific to each clinical situation because the accuracy (sensitivity and specificity) of a test is likely to be influenced by <u>factors including</u> “the time from onset of infection, the concentration of virus in the specimen, the quality and processing of the specimen collected from a person, and the precise formulation of the reagents in the test kits.”^21 22^. The distribution of test statistics can be both individual specific and also identical and independent across test repetitions. Note, moreover, that results for an individual are likely to be correlated over test repetitions, even though the tests are probabilistically independent. For instance, a patient with a high viral load may be very likely to have a positive test result in each round, which quite appropriately leads to serial correlation in test results. But as long as the likelihood of a false negative is constant across rounds then the assumption of an identical and independent distribution holds^3^. Whether the assumption of distributions that are identical and independent across repetitions is correct can be empirically investigated by determining whether the likelihood of false positives and false negatives appears constant across rounds under test repetition. We are not aware of any study examining this question in the present context. It should therefore be the subject of future research.

Note that *cost of test 3 increases arithmetically but sensitivity increases non-linearly with the number of repetitions, asymptotically approaching the upper bound of 1*. As a result, if c2 is sufficiently low relative to c1, then there is always an M, call it M*, such that p3(M*) = 1-(1-p2)^M*^ > p1 and M*c2 < c1. In other words, if c2/c1 is sufficiently low, then test 3 (M) is cheaper than test 1 and more sensitive. In this simple exposition, we have not taken in to account other concerns relevant to test comparisons, such as the consequences of delays in test processing, and the clinical relevance of distinct tests as a result of their different LODs or other considerations.

The gain in sensitivity in the example above is “purchased” at the expense of decreased specificity. If the specificity of Test 2 is q2 then the specificity of Test 3(M) based on repetition is q3(M) = (q2)^M^. But note – crucially -that if q2 is high to begin with then the loss in specificity due to repetition will be small. For instance, if specificity is 99.5% to begin with then repetition twice lowers specificity to a level just above 99%. In the case of antigen tests for Covid-19, which have very high specificity, the loss in specificity due to repetition is therefore negligible. In any case, repeat testing can result in improvements in *both* specificity and sensitivity if additional repetitions are undertaken and if more general decision rules than the “single positive counts as overall positive” rule considered in the example above are applied to interpreting the ensemble of test results. (See Appendix 2 for reasoning and Table 1 below for numerical results).

**Table 1:** Sensitivity and Specificity for different PRT designs and the BinaxNOW antigen test.

| n | k |  |  |  |  |
| --- | --- | --- | --- | --- | --- |
|  | 1 | 2 | 3 | 4 | 5 |
| 1 | 0.93 |  |  |  |  |
| 2 | 1.00 | 0.86 |  |  |  |
| 3 | 1.00 | 0.99 | 0.80 |  |  |
| 4 | 1.00 | 1.00 | 0.97 | 0.75 |  |
| 5 | 1.00 | 1.00 | 1.00 | 0.96 | 0.70 |

**SPECIFICITY**
| n | k |  |  |  |  |
| --- | --- | --- | --- | --- | --- |
|  | 1 | 2 | 3 | 4 | 5 |
| 1 | 1.00 |  |  |  |  |
| 2 | 0.99 | 1.00 |  |  |  |
| 3 | 0.99 | 1.00 | 1.00 |  |  |
| 4 | 0.98 | 1.00 | 1.00 | 1.00 |  |
| 5 | 0.98 | 1.00 | 1.00 | 1.00 | 1.00 |

The statistical reasoning underlying PRT is straightforward. The repetition of a test provides more information and can therefore only aid inference. But whether it improves one criterion (sensitivity) at the expense of another (specificity), or whether it improves both, depends on how the information gained is distributed across the distinct objectives, and this in turn depends on the specific decision rule applied to interpret the ensemble of test results.

Apply the reasoning above to the Abbott BinaxNOW COVID-19 rapid antigen test, adopting specific parameter assumptions corresponding to the empirical characteristics of this test reported above. Namely for the antigen test, c2 = $5^23^, sensitivity p2 = 0.93 and specificity q2^24^=0.995. For the PCR test suppose that <u>c1 = $50</u>^25^ sensitivity p1=0.98, specificity q1=0.99. These values of sensitivity and specificity may be above what is clinically achieved for PCR tests^26^; notably if one considers infectivity in viral cultures as a measure of ‘clinically relevant disease positivity’, then the specificity and notably the accuracy (positive predictive value) of the PCR test may be considerably lower^3,15,27^. However, the exact numerical assumptions for antigen and PCR test sensitivity and specificity made here do not influence the larger point. They can be modified as desired, since these values may depend, as noted above, on the site and quality of sample collection, among other factors^28^. Recent studies summarize estimates of sensitivity and specificity for other rapid antigen tests, and can be used as a basis for alternative calculations if desired^24 22^. The effect of parallel repeat testing on increasing sensitivity for different assumptions concerning the underlying test is shown in the annex spreadsheet (first tab), downloadable <u>here</u>: https://tinyurl.com/46tu9nre. As can be seen, the greatest proportional gains in sensitivity are for the lowest initial levels, in the range described.

Repeating Test 2 (corresponding to the BinaxNow test) twice, and considering the test subject to be positive if he or she is positive in any single round, leads to a test procedure (Test 3) that will have a cost of $10, a sensitivity of 0.99>p1, and specificity of 0.99, only marginally reduced from its original level, and in this example the same as the specificity of a PCR test. For a cost of $10, one-fifth that of the “gold standard” PCR test, one can achieve *superior* results than the PCR test, for the assumptions about that test made above. Moreover, the antigen tests used are widely available and give rapid results, unlike PCR tests in many cases.

With three repetitions and considering the test subject to be positive if he or she is positive in any single round, sensitivity p3(3) is almost 1 and specificity is only marginally reduced to a little less than 0.99. There is therefore a significant benefit in terms of increase in sensitivity with a marginal decrease in specificity, maintaining an extremely high level of specificity overall. For sensitivity and specificity corresponding to the BinaxNOW Antigen test, such a composite test can improve both specificity and sensitivity of the underlying test for an individual, raising both to the level of greater than 99 percent. In this case, given the relatively low cost of antigen testing, it is compelling to perform a composite test based on repetition. In the next section, we show that with wider decision rules, there need not be any tradeoff at all, and both sensitivity and specificity of a test can be improved together.

## III. Improving both sensitivity and specificity

By applying a more sophisticated decision rule for interpreting the ensemble of results, both sensitivity and specificity of the test being applied to an individual can be improved. For instance, the overall determination of whether a composite test is positive can be based on the number of positive antigen test results in an ensemble for a given number of repetitions, e.g. at least two out of three, rather than a single positive test. In particular, using the same parameters assumed above, corresponding to the BinaxNOW antigen test, sensitivity would be improved from 0.93 to almost 0.99 and specificity from 0.995 to almost 1 (0.999925) by requiring at least two positives out of three test results rather than a single positive in order to arrive at an overall positive determination (see Appendix 2 for reasoning and Table 1 below for numerical estimates). Consider similarly the case in which least two out of four tests in an ensemble being positive, rather than a single positive test, suffices to trigger an overall positive determination. Both specificity and sensitivity can be improved in this case to values greater than 0.999. Repeating an antigen test four times and analyzing the results in parallel provides a way to markedly improve both sensitivity and specificity.

Table 1 describes the levels of sensitivity and specificity of a composite test that will result from different possible PRT designs (with n repetitions, and at least k of n underlying tests being positive sufficing for an overall positive determination) for parameters corresponding to the BinaxNOW antigen test (specifically sensitivity of 0.93 and specificity of 0.995).

## IV. The population-level consequences of Parallel Rapid Testing

Until now we have considered the impact of Parallel Rapid Testing on the sensitivity and specificity of tests applied to individuals. A distinct issue concerns the population-level consequences of applying a particular testing approach. These may be important from two points of view. The prevalence of a disease in the population being tested can affect (a) how informative a test should be taken to be for a specific individual in that population, and (b) how successfully a test contributes to identification of individuals in the population whose contacts should be limited. These two perspectives are respectively of relevance to clinical management and public health strategies. We discuss these in turn below, assuming for simplicity that a test has the same levels of sensitivity and specificity when applied to all individuals in the population.

### (a) Population Prevalence and Clinical Relevance

As is well known, the positive and negative predictive values of a test, describing how likely an individual who tests positive is actually to be positive, or who tests negative is actually to be negative, depend on the proportion of a population that actually has the condition being tested (i.e. is a “true” positive). An increase in both sensitivity and specificity, achieved by applying a rule of the kind described in Section III (for instance, n=3, k=2) will have an unambiguously beneficial effect, raising both positive and negative predictive values, and lowering the proportions of false positives and false negatives among all those tested. This is not the case, however, if a rule of the kind discussed in Section II is applied (for instance n=2, k=1) which raises sensitivity but lowers specificity. An increase in sensitivity of a test applied to a population will lower the number of false negatives and raise the number of true positives and a decrease in specificity will lower the number of true negatives and raise the number of false positives. As the negative predictive value is the number of true negatives divided by the total number of negatives, the net impact is a priori ambiguous, and depends on the magnitude of each of these effects and on the affected population fractions. As the positive predictive value is the number of true positives divided by the total number of positives, the net impact on it too is ambiguous and similarly would depend on the magnitude of the effects and the affected population fractions (see Appendix Three).

As we show in the annex spreadsheet (third tab) for the parameters we have considered above and some plausible infected population shares (15%, 10%, and 5%) the use of parallel rapid antigen tests (n=2, k=1) raises negative predictive values and lowers positive predictive values as compared to the single test case. Even so, the positive and negative predictive values remain above those of PCR tests, for the assumed values of sensitivity and specificity of the latter. As we show in the annex spreadsheet (fourth tab) for the parameters we have considered above, and some plausible infected population shares (15%, 10%, and 5%) the use of parallel rapid antigen tests (n=3, k=2) raises both negative predictive values and positive predictive values as compared to the single test case. Both the positive and negative predictive values rise above the values for PCR tests, given the assumed values of sensitivity and specificity of the latter, and indeed approach 100%.

### (b) Population Prevalence and Public Health Strategies

The likelihood of false negatives among all persons tested is of direct relevance for assessing alternative public health strategies, as it will determine the efficacy of testing as a strategy for containing or suppressing the disease. The likelihood of false positives among all persons tested is relevant for assessing public health strategies too, as it will shape the cost to society arising from unnecessary restrictions of contacts. As we show in the annex spreadsheet (third tab) for the parameters we have considered above and some plausible infected population shares (15%, 10%, and 5%) the use of parallel rapid antigen tests (n=2, k=1) raises the percentage of false positives in the tested population and lowers the percentage of false negatives in the tested population as compared to the single test case. Even so, these values are above those of PCR tests, for the assumed values of sensitivity and specificity of the latter. As we show in the annex spreadsheet (fourth tab) for the parameters we have considered above, and some plausible shares of the tested population assumed infected (15%, 10%, and 5%) the use of rapid antigen tests (n=3, k=2) lowers both the share of false positives and false negatives in the tested population. In both cases, these values are below those for PCR tests (less than 0.1% for false negatives and less than 1% for false positives), given the assumed values of sensitivity and specificity. This assessment assumes that the testing strategy homogenously applying tests of a single kind (e.g. all PCR tests or all parallel rapid antigen tests).

## V. Other considerations in determining the composite test design

The number of repetitions and the appropriate number of positive results in an ensemble to be applied as a threshold should be identified based on suitable decision criteria, for instance concerning the costs of testing and the potential implications for disease control. The implications of different decision rules for disease transmission should be studied in the context of epidemiological models considering possible dynamics.

In practice, composite tests may be attractive not only because of their statistical properties, which may make them comparable or superior to more expensive alternatives, but because using them may allow the excess demand for more “sophisticated” tests (i.e. PCR tests) to be reduced. Focusing PCR tests on those who test positive in one round (or more) of the parallel rapid test may result in many fewer demands on PCR testing. If test processing capacity is constrained, this may reduce lags in processing tests, and therefore ensure greater societal benefit from the testing that does take place.

Even Parallel Rapid Testing using antigen test) might be limited to those we have specific reason to believe are at risk of receiving the disease, of being contagious or of having severe adverse outcomes. Parallel Rapid Testing could also be restricted to sub-populations which have tested positive during pooled PCR testing or where the prevalence of the disease among those tested reaches a specific threshold (e.g. > 5% ^22^) for restriction of activities). The most desirable approach will depend on risk assessment. A targeted approach focusing testing on specific individuals should be shaped by information about risks of disease transmission and outcomes, e.g. if it is thought that <u>symptomatic transmission of the disease is most important</u> or that it is most likely to be spread among certain groups (see previously mentioned CDC guidance). Parallel Rapid Testing addresses a different problem than does sequential testing. The latter may provide important information in the scenario corresponding to an initial test at time T1 (Figure 1), which may fail to detect an incipient infection. As outlined in Appendix 1, it remains challenging to form correct inferences about the true level of viral concentration in some individuals due to changes in viral load over time. Nonetheless, sequential testing within a short time period (e.g. consecutive or alternate days) is a useful complementary strategy, especially when community transmission and prevalence rates are higher, or in particularly vulnerable populations (e.g. nursing home populations). In fact, parallel and sequential testing can be combined, so as to offer a more accurate as well as continuous picture of the disease status of individuals. While parallel repeated tests aid inference about whether a patient currently possesses a clinically relevant level of the virus, they cannot tell us whether the patient will do so in the future, as the level of viral load in the patient at the time a test is taken depends on the point in the course of an infection that has been reached. For this reason, the two approaches to repetition -parallel and serial -are complementary. Additional tests at a given time increase the confidence with which inferences can be made about the situation right then, whereas additional tests taken over time help to capture how the infection status of the individual evolves.

With further reductions in the cost of antigen testing, we can imagine widespread use of Parallel Rapid Tests as a basis for making nearly immediate determinations of who may be permitted to participate in societal activities. This possibility should at least be recognized, without antigen testing being viewed as always and everywhere inferior to PCR testing for the purpose of controlling the spread of COVID-19 in the community, which – as we have just shown – it is not.

## Data Availability

No data were used.

https://reddytoread.files.wordpress.com/2021/03/testrepetitions19march2021.xlsx

## Acknowledgements

We would like to thank David K. Miles, Arnab Acharya, Jishnu Das, Sanmay Das, Duncan Foley, Paulo dos Santos, David K. Miles and Robert Rowthorn for their helpful suggestions.

## Supplemental Material

### APPENDIX 1: Inferences under Parallel vs. Sequential Testing

A more familiar alternative to parallel testing is sequential testing, in which tests are conducted for the same individual at distinct times (the sense in which “repeat” testing is referred to in <u>current CDC recommendations</u>)^29^. What are the advantages or disadvantages of sequential as compared to parallel testing? Specifically, assuming that more tests always give more information that is potentially useful in inference (e.g. as to whether an individual possesses the disease, or is infective, at a given time) how can this information be used in the case of sequential testing, and how does this compare to the application of information from multiple tests in parallel testing?

Suppose that additional tests are conducted at n-1 different times after the initial test, s_1_,s_2_,..s_n-1_ where s_0_=0 and s_1_,..s_n-1_ refer to lengths of time after the initial test. Note that if all of the n tests are conducted in a single moment, s_1_=s_2_=..=s_n-1_=0, then the sequential testing case collapses to the parallel testing case. In this special case, since there is no reason to treat one test as carrying more information than another, the weight attached in inference to the various tests must be the same. But if the times are not identical then the question arises of what weight to give to tests undertaken at different times in order to form inferences about the current situation (in particular whether or not the individual tested should be deemed to possess the disease, to be potentially infectious, etc.).

A series of tests could have very different implications for inference about present status depending on when the tests are undertaken. A test conducted in the near future (e.g. in the next few days) may help to resolve whether an individual presently has the disease, or is infectious. It is reasonable to presume that greater weight should be attached to more recent tests in determining an individual’s current status^4^. But how exactly should the weight attached to different tests vary with time, when making an inference about current disease status, given the possibility of error in any one test, including the most recent one?

Suppose that each test conducted s_i_ units of time after the initial test is associated with a result R_i_ which can take on one of two possible values, P or N, for positive or negative. In that case the object of inference is to assign an individual an overall P or N describing a judgment about *current* status based upon a given ordered set of results <u>R</u> = {R_i_} corresponding to the tests administered, initially and at various subsequent times, <u>s</u>={s_i_}.

Suppose that an individual is considered a true positive if the virus is present with a measure of viral load, v, greater than a given clinically relevant threshold, z. The choice of threshold necessarily depends on the clinical or public health purpose: e.g. determining particular risks an individual patient may face that are conditional on infection status, identifying the appropriate individual treatment protocol, deciding whether the patient should be isolated in the interests of preventing spread of the infection etc. Although the interpretation of this purpose does not influence our formal analysis, for purposes of exposition we have assumed that the main concern influencing the choice of threshold is infectivity^5^.

Suppose further that the course of the infection is one in which the measure of the presence of the virus changes over time according to a function F(t) describing viral load; where t describes the length of time that has elapsed from the onset of exposure to the virus, possibly varying across individuals. An individual is a true positive if and only if F(t) ≥ z. It is, however, unknown if or when an individual has been “infected” and if so, where in the course of the infection the individual has been tested. If this information were known, it would be possible, on the basis of F(t) and z, to determine the expected sequence of test results, <u>R</u>, that would arise from an error-free test. But it is precisely because this information is not known that testing is needed. Unfortunately, interpreting the individual’s present status on the basis of sequential test results, when it is unknown whether the individual is infected at all, and if so how far the infection has progressed, poses a challenging inferential problem, compounded by the presence of test errors.

To see why, consider tests administered at the time of an initial test, t=t_0_, and subsequently, with <u>s</u>={0,24,48} and the units taken to be hours. In other words, suppose there were three tests; one at an initial time at an unknown point in the course of an infection, and one each 24 and 48 hours after it. The individual possesses viral concentrations at levels F(t), F(t+24) and F(t+48). Because the level of the viral load F(t+h) varies with t and h for an infected individual, and in particular whether the viral load is above or below the threshold z depends on this, the results that would be returned even by an error-free test at each of these times also varies accordingly. In principle, various possible sequences of test results could all be consistent with the individual being a “true positive” at the time of the latest test, even for a perfectly accurate test, and test error increases the range of possibilities even further.

This is illustrated by the diagram in Figure 1, with a curve describing viral load as a function of time, F(t), ascending to above the threshold, z, and then descending beneath it, as the course of an infection proceeds. Consider two tests taken at an initial time T1 (described by the right hand most solid bar), and at later time T2. A perfectly accurate test, where a true positive is defined by the threshold of clinical relevance, would return results {N,N} respectively for the tests. But what if the point in the course of the infection reached in the present moment were instead depicted by the righthand most dashed bar, Δ units of time later in the course of the infection than the initial test considered earlier. In that case, a perfectly accurate test, relative to the threshold of clinical relevance, would return results {P,P} respectively for the tests. With different starting positions, and lengths of time between tests, other possibilities might arise including {N,P} and {P,N},}. When the possibility of inaccuracy of a test is introduced, along with a possible distinction between the threshold of clinical relevance and the LOD, the difficulty of inferring the current situation correctly is compounded. One may improve on the accuracy of sequential testing by spacing the tests appropriately, informed by knowledge of the dynamics of viral replication; however, the rise of newer variants with unknown kinetics of viral replication may make determination of the appropriate time interval between testing harder to ascertain.

Assessing whether a given sequence of test results suggests a specific conclusion about the present situation (e.g. that the true condition of the individual is “positive”) is challenging. Although some sequences will be more likely to indicate that the individual is currently truly infected than others, it is not obvious how to determine the associated probability exactly. The likelihood with which a set of test results indicates that the individual is a “true positive” or a “true negative” will depend not only on the sensitivity and specificity of the test but on various other factors, including the likelihood that an individual in the tested population is infected; the expected course of an infection for an individual, F; the likelihood that the individual is at a specific point in the course of the infection, t; the lengths of time separating the present test from prior tests, s_i_; the threshold of clinical relevance for determining a positive result, z, if distinct from the LOD; the likelihood of false positives and negatives at each point in the course of the infection (which may well vary with t and z, as well as with the test applied). The range of factors involved can make it very difficult to determine, based on sequential test results, the likelihood that the individual is currently a “true positive” or a “true negative”.

More frequent tests conducted closer to the present are likely to be more informative about the present situation -in the limiting case, approaching parallel testing. For example, one or more positive results arising in a short period are likely to increase the chance that an individual is currently positive. We are not aware of a complete theory of how to employ information from a sequence of tests to draw inferences about the present disease status of an individual. The application of such a theory would in any case depend on facts (such as the function F that represents the typical course of the infection for an individual) which remain little understood in the case of Covid-19.

### APPENDIX 2: Combinatorial Mathematics of Parallel Rapid Testing

How do the sensitivity and the specificity of a composite test relate in general to those of the underlying test? Can we understand what designs of a composite test cause specificity to be gained at the expense of losing specificity, and what designs cause these test properties to be improved jointly?

Consider a protocol in which a “composite test”, *T*^, based on an underlying test is repeated n times. The underlying test returns a value of positive or negative. Suppose that the composite test is considered to be positive if a critical threshold is reached in which at least 1 ≤ k ≤ n of the underlying tests are positive. We will refer to the composite test based on n repetitions of an underlying test and a critical threshold k by *T*^(n, k).

The case *T*^(n, 1) is that of an “OR” composite test, in which a positive test in *any* of the underlying positive tests suffices for a positive result in the composite test. The case *T*^(n, n) is that of an “AND” composite test, in which a positive test in *all* of the underlying positive tests is necessary (and sufficient) for a positive result in the composite test. The “in between” cases are ones in which having some number of positive tests (more than one but not all) in the ensemble of tests is necessary and sufficient for a positive result in the composite test.

If the sensitivity of an underlying test is p then the proportion of false negatives associated with it is (1-p). It is straightforward to deduce, because whether a false negative occurs or not is a binary outcome, that a binomial distribution applies – making, as above, an assumption of the identicality and independence of the distribution of test results across repetitions.

The chance of *exactly i* false negatives arising out of *n* repetitions of the underlying test is 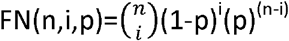. The chance of a false negative arising in the composite test, *T*^(n, k), as a whole, due to one arising in n-k+ 1 or more tests in the ensemble of tests (thereby making an overall positive assessment in the composite test, arising from *k* or more positive test results, impossible), is therefore 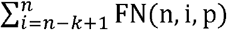. The sensitivity of the composite test is then 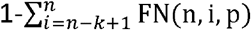.

Similarly, if the specificity of an underlying test is q then the proportion of false positives associated with it is (1-q). The chance of *exactly i* false positives arising out of *n* repetitions of the underlying test is 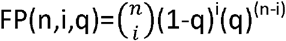. The chance of a false positive arising in the composite test, *T*^(n, k), as a whole, due to one arising in *k* or more tests in the ensemble of tests (thereby making an overall negative assessment in the composite test impossible), is therefore 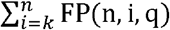. The specificity of the composite test is then 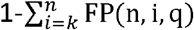.

Is there (n, k) such that the sensitivity and specificity of *T*^(n, k) are both greater than for the underlying test? The answer is yes. Fixing the number of tests n and increasing (decreasing) the threshold k leads to a gain (loss) in specificity at the cost of a loss (gain) in sensitivity, with minimum sensitivity being attained at k=n and maximum sensitivity being attained at k=0. However, increasing n and choosing k appropriately so as to “distribute” the additional information collected between the two objectives of reducing false positives and reducing false negatives (moving “down” rows while avoiding the extreme right and left of Pascal’s Triangle) can, at least for suitable values of p and q, lead to an improvement in both sensitivity and specificity. This can be checked algebraically or computationally. In principle, mixed strategies involving applying different thresholds with some probability makes possible additional levels of sensitivity and specificity between the values corresponding to integer values of k.

Let us verify the possibility of increasing both sensitivity and specificity at the same time, through repeat testing, with some examples.

Case (n=3, k=2): If is straightforward to show algebraically using the expressions given above that if sensitivity of the underlying test, p, is greater than one half and its specificity, q, is greater than one half then applying the rule that at least two out of three “positives” in the underlying test makes for a positive in the composite test will lead to an improvement in both. Whether a parallel repeated test involves a tradeoff between sensitivity and specificity or a potential gain all around is therefore in part an empirical question involving the properties of the underlying test to be repeated. Sensitivity of the composite test is given by [1-((1-p)^3^+3(1-p)^2^p)] and specificity is given by [1-((1-q)^3^+3(1-q)^2^q)]. Therefore, the absolute gain in sensitivity is [-(1-p)^3^ -3(1-p)^2^p-p+1] and the absolute gain in specificity is [–[(1-q)^3^-3(1-q)^2^q-q+1] as a result of using the composite test rather than the unrepeated underlying test. The absolute gain in turn increases and then decreases, in both cases, with the sensitivity and specificity of the underlying test (reaching a common peak at p=q=0.79).

Case (n=4, k=2): Similarly, if sensitivity of the underlying test, p is greater than 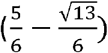, or around 0.23, then applying the rule that at least two out of four “positives” in the underlying test makes for a positive in the composite test will lead to an improvement in sensitivity. The sensitivity in this case is [1-((1-p)^4^+4(1-p)^3^p)] and the absolute gain in sensitivity is therefore given by [-(1-p)^4^-4(1-p)^3^p-p+1]. If specificity of the underlying test, q is greater than 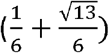, or around 0.77, then applying the rule that at least two out of four “positives” in the underlying test makes for a positive in the composite test will lead to an improvement in specificity. The specificity is [1-((1-q)^4^+4(1-q)^3^q) +6(1-q)^2^q^2^)] and the absolute gain in specificity is therefore given by [-(1-q)^4^-4(1-q)^3^q-6(1-q)^2^q^2^-q+1]. It may be checked that, in this case too, both the absolute gain in sensitivity and the absolute gain in specificity as a result of using the composite test rather than the (unrepeated) underlying test increase and then decrease with the sensitivity and specificity of the underlying test, once they rise above the critical thresholds for the composite test to result in an improvement at all.

Case (n=4, k=3): We get “flipped” results when we increase k to 3, which is not surprising given the symmetric nature of the binomial distribution. If sensitivity of the underlying test, p, is greater than around 0.77 then applying the rule that at least three out of four “positives” in the underlying test makes for a positive in the composite test will lead to an improvement in sensitivity. The sensitivity in this case is [1-((1-p)^4^+4(1-p)^3^p+6(1-p)^2^p^2^)] and the absolute gain in sensitivity is therefore given by [-(1-p)^4^-4(1-p)^3^p-6(1-p)^2^p^2^-p+1]. If specificity of the underlying test, q, is greater than around 0.23, then applying the rule that at least two out of four “positives” in the underlying test makes for a positive in the composite test will lead to an improvement in specificity. The specificity is [1-((1-q)^4^+4(1-q)^3^q)] and the absolute gain in specificity is therefore given by [-(1-q)^4^-4(1-q)^3^q-q+1].Again, both the absolute gain in sensitivity and the absolute gain in specificity as a result of using the composite test rather than the (unrepeated) underlying test increase and then decrease with the sensitivity and specificity of the underlying test, once they rise above the critical thresholds for the composite test to result in an improvement at all.

The annex spreadsheet we provide (second tab) offers an easy way to test the impact of different assumptions about sensitivity and specificity of the tests being compared based on properties of the underlying tests (p1, p2, q1, q2) and composite test designs (n,k) for the parallel repeated test.

### APPENDIX 3: Impact of Simultaneous Changes in Test Sensitivity and Specificity on Positive and Negative Predictive Values

As previously, let p represent sensitivity and q represent specificity. Let *θ* represent prevalence of the infection in the tested population as judged by criteria of clinical relevance. In that case positive and negative predictive values are represented respectively by:

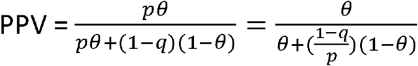

and

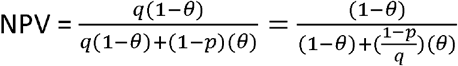

If p rises ((1-p) falls) and q falls ((1-q) rises)) then whether 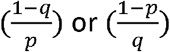 respectively increase or decrease depends on the relative magnitude of the changes in p and q. If p and q both rise then this causes an unambiguous improvement in both PPV and NPV. The population prevalence does not directly determine the sign of the change in PPV and NPV but it does affect its magnitude. It can also be checked that the proportion of false positives among all those tested (1 -*p θ*) falls (rises) as sensitivity increases (decreases) whereas the proportion of false negatives among all those tested (1-q(1 -*θ*)) falls (rises) as specificity increases (decreases).

## Footnotes

2 We would like to thank David K. Miles, Arnab Acharya, Jishnu Das, Sanmay Das, Duncan Foley, Paulo dos Santos, David K. Miles and Robert Rowthorn for their helpful suggestions.

3 The independence requirement is that the distribution of test statistics (i.e. the likelihood of false positives or negatives) in any given round is not influenced by the results of prior rounds. We do not judge a violation of independence to be likely, as it is hard to envision how early test results, especially if not revealed to the operator prior to subsequent tests, would influence the statistical properties of later tests. Even if independence holds, it is possible that the assumption of identical distributions does not hold, for example due to operator fatigue or other factors which cause later rounds to have a systematically different level of accuracy than earlier rounds. The assumption can be relaxed if empirical information becomes available that suggests that the probability of a false positive or false negative changes across test repetitions to a degree that is inappropriate to ignore. The calculations for the sensitivity and specificity of parallel repeated tests would have to be made differently in such a case, assuming independence but not identicality, but are conceptually straightforward as long as the probabilities specific to each round are known.

4 If tests are considered to be informative about previous occurrences of an illness as well as current occurrences, this may generate greater complexity. For instance, tests conducted in the distant past may have relevance to determining whether an individual presently has the disease, or is infectious, if it is thought that the individual may have acquired immunity as a result of a past occurrence. This issue is disregarded here, in order to focus on clinical assessment protocols involving tests undertaken during or in proximity to a current disease episode.

5 It is important to note that most studies of test properties do not appear to recognize that assessments of sensitivity and specificity cannot be made in abstraction from specific clinical or public health objectives.

